# “Big Events” and HIV transmission dynamics: estimating time since HIV infection from deep sequencing data among sex workers and their clients in Dnipro, Ukraine

**DOI:** 10.1101/2025.06.13.25329586

**Authors:** François Cholette, Nicole Herpai, Leigh M. McClarty, Olga Balakireva, Daria Pavlova, Anna Lopatenko, Rupert Capiña, Paul Sandstrom, Michael Pickles, Evelyn Forget, Sharmistha Mishra, Marissa L. Becker, the Dynamics Study

**Author notes:** Correspondence: François Cholette, National Sexually Transmitted and Blood-Borne Infections Laboratories, National Microbiology Laboratory, Public Health Agency of Canada, 745 Logan Avenue, Winnipeg, Manitoba R3E 3L5, Canada. In memoriam (Olga Balakireva). These authors contributed equally to this work (François Cholette, Nicole Herpai, Leigh M. McClarty). These senior authors contributed equally to this work (Sharmistha Mishra, Marissa L. Becker). Membership of the *Dynamics* Study Team is provided in the Acknowledgments.

## Abstract

**Background:** Major geopolitical events and structural shocks are thought to play a significant role in shaping HIV epidemics by influencing individual behaviours, reshaping social networks, and impacting HIV prevention and treatment programs. Here, we describe individual-level measures of estimated time since HIV infection (ETI) from viral next generation sequencing data among female sex workers and their clients in relation to significant geopolitical events in Ukraine.

**Methods:** The *Dynamics study*, is a cross-sectional integrated biological and behavioural survey conducted among female sex workers and their clients in Dnipro, Ukraine (December 2017 to March 2018). We were able to successfully sequence a portion of the HIV *pol* gene on dried blood spot specimens among *n* = 5/9 clients and *n* = 5/16 female sex workers who tested positive for HIV (total *n* = 10/25) using an in-house drug resistance genotyping assay. The “HIV EVO” Intrapatient HIV Evolution web-based tool (https://biozentrum.unibas.ch/) was used to infer ETI from viral diversity.

**Results:** The median ETI for female sex workers and their clients was 5.4 years (IQR = 2.9, 6.6) and 6.5 years (IQR = 5.4, 10.8) respectively. Nearly all HIV acquisition events (*n*=7/10; 70%) were estimated to have occurred between the Great Recession (2008 – 2009) and the War in Donbas (May 2014 – February 2022). In general, ETI suggests that HIV acquisition occurred earlier among clients (2012 [IQR = 2007, 2013]) compared to sex workers (2013 [IQR = 2012, 2016]).

**Conclusion:** Our findings suggest that most HIV acquisition in this small subset of female sex workers and clients living with HIV, occurred during periods of economic decline. Molecular studies on timing of HIV acquisition against timing of major geopolitical events offer a novel way to contextualize how such events may shape transmission patterns.

## INTRODUCTION

The HIV epidemic in Ukraine is considered one of the most significant in Europe ^1^, with an estimated 240,000 people living with HIV ^2^ in the country. A growing number of infections stem from unmet HIV prevention and treatment needs of female sex workers (FSW) and their clients ^3^. It is estimated that 3.1% of Ukraine’s approximate 86,000 FSW are living with HIV ^2^, while prevalence estimates for clients of FSW range between 2% and 23% ^1 3-5^.

Major geopolitical events and structural shocks, termed “big events”, such as wars, economic transitions, global pandemics, and political uprisings are theorized to play a significant role in shaping HIV epidemics by influencing individual behaviours, re-shaping social network structures, and impacting HIV prevention and treatment programs^6^. A series of big events including the Revolution of Dignity, illegal Russian annexation of Crimea, invasion of the eastern region of Donbas followed by recent full-scale war, jeopardizes the progress made towards curbing the HIV epidemic in Ukraine ^3^. For example, Vasylyeva and colleagues have demonstrated that the annexation of Crimea and Donbas conflict impacted the spread of HIV in Ukraine, specifically on the patterns of viral dissemination ^7^.

Here, we describe individual-level measures of estimated time since HIV infection (ETI), defined as the estimated number of years prior to sampling when HIV acquisition likely occurred, inferred from viral next generation sequencing (NGS) data ^8^. These estimates are then juxtaposed on a timeline of major geopolitical events in Ukraine. Our approach, which overlays two sources of seemingly disparate data (intra-host viral diversity and geopolitical events) provides opportunities to re-examine conceptual and future analytical insights into structural drivers of HIV transmission and acquisition.

## MATERIALS AND METHODS

### Study setting

The *Dynamics* Study was a cross-sectional behavioural and biological survey of FSW and their clients in Dnipro, Ukraine, conducted from 10 October 2017 through 11 February 2018 ^4^. Study participants were identified in validated sex work hotspots and invited to participate in the study by outreach workers. Consenting participants completed an interviewer-facilitated behavioural survey, underwent HIV rapid testing, and provided a dried blood spot (DBS) sample for confirmatory serological testing (Avioq HIV Microelisa System, Avioq Inc., Durham, United States) and viral genomic sequencing at the National HIV and Retrovirology Laboratory (Public Health Agency of Canada, Winnipeg, Canada). Pre- and post-test counselling was provided according to Ukrainian national guidelines.

### HIV *pol* sequencing

We attempted to sequence a portion of the HIV *pol* gene (position 2,074 – 3,334 on HXB2, accession no. K03455) on all HIV seropositive DBS specimens using a validated, in-house, drug resistance mutation genotyping assay, described elsewhere ^9 10^. Sequencing libraries were prepared using the Nextera XT DNA library preparation kit (Illumina, San Diego, United States). All steps of the library preparation were performed using an epMotion 5075t liquid handling station (Eppendorf, Hamburg, Germany). Sequencing was performed on a MiSeq platform (Illumina) using v2 MiSeq reagent kits (300 cycles; Illumina) according to the manufacturer’s instructions. MiSeq reads were reference mapped to HIV HXB2 (accession no. K03455) using HyDRA Web (http://hydra.canada.ca).

### Estimation of time since infection

Hamming distance and the HIV EVO Intrapatient HIV Evolution web-based tool (https://hiv.biozentrum.unibas.ch/) were used to infer time since HIV infection (ETI) from the diversity of NGS reads ^8^.

### Descriptive analysis

*Dynamics* study participants’ sociodemographic characteristics are reported using descriptive statistics. Continuous data are summarized using the median and interquartile range (IQR).

Categorical data are presented using exact numbers and proportions. All analyses were conducted using SPSS Statistics v28 (IBM, Armonk, United States). The figure was generated using Python 3.10.7 and Matplotlib v3.6.0 ^11^.

## RESULTS

A total of 370 clients (88.9%) and 560 FSWs (86.0%) consented to complete the behavioural survey. All participants except one provided (99.9%) a DBS sample. Serological testing identified 25 (2.7%) participants living with HIV, of whom 9 were clients and 16 were sex workers. We successfully sequenced HIV *pol* from 10 (*n* = 5/9 clients and *n* = 5/16 sex workers) seropositive DBS specimens. All sequences were classified as subtype A (sub-subtype A6) according to COMET ^12 13^.

The median estimated time of HIV infection (ETI) for clients and sex workers was found to be 6.5 years (IQR = 5.4, 10.8) and 5.4 years (IQR = 2.9, 6.6) prior to DBS sample collection, respectively. ETI was then compared to the DBS sample collection date to infer the date of HIV acquisition (Fig. 1). Seven of the 10 HIV acquisitions occurred between the Great Recession (2008/2009) and the beginning of war in Donbas (2014). The pattern of ETI suggests infections among clients occurred earlier (median of 2012 [IQR = 2007, 2013]) than those among sex workers (2013 [IQR = 2012, 2016]). However, clients were older (median age = 37 years; IQR = 31, 43) in comparison to sex workers (median age = 30 years; IQR = 26, 44).

**Figure 1.**
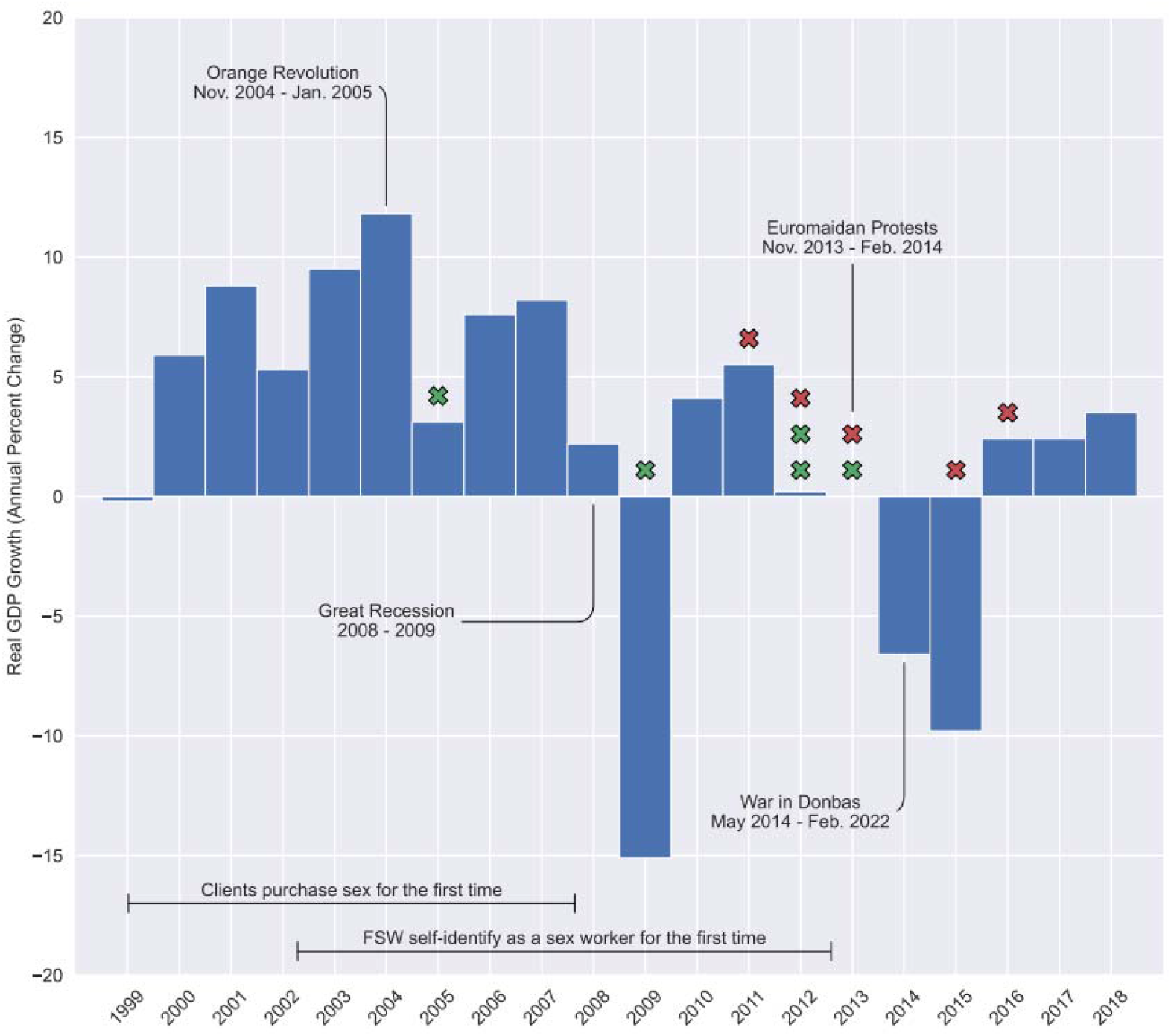
Estimated time since HIV infection among female sex workers (red “X”) and clients (green “X”) in Dnipro, Ukraine relative to recent political, economic, and social events. Real GDP growth data retrieved from The World Bank (https://data.worldbank.org/indicator/NY.GDP.MKTP.KD.ZG?locations=UA).

## DISCUSSION

This exploratory analysis highlights the potential influence of structural shocks on HIV acquisition among sex workers and their clients in Dnipro, Ukraine. Using viral NGS data, we estimated individual-level estimates of time since HIV infection and situated these infections within a timeline of major geopolitical events in Ukraine. Among the ten successfully sequenced samples—five from sex workers and five from clients—the majority of estimated HIV acquisitions occurred between 2008 and 2014, following the Great Recession and prior to the onset of the Donbas conflict. Estimated dates of infection were earlier among clients than sex workers, which may partly reflect age-related differences, as clients were typically older. These findings suggest that periods of economic instability may have shaped patterns of HIV transmission in this population, stressing the importance of contextualizing molecular data within broader structural and temporal frameworks.

Major structural shocks and geopolitical events such as wars, economic recessions, global pandemics, and climate change can have a significant impact on sex work and the people involved in it ^14-18^. Elmes and colleagues (2017) illustrate how sex work was reorganized in Zimbabwe amidst a shrinking economy. Female sex workers adapted by changing the locality of sex negotiations, strategies for attracting clients, and payment systems ^15^. Similarly, social and political unrest have impacted *Dynamics* Study participants. Most female sex workers (80%) surveyed during the *Dynamics* Study financially supported at least one dependent and some described providing support for family members as a key motivator for entering sex work, especially in times of economic uncertainty ^19^. Among those already engaged in sex work, economic downturns likely contributed to reduced client volumes and diminished disposable income among clients, as suggested by a qualitative analysis by Lazarus et al^20^. In response to these financial pressures, some sex workers may have accepted higher-risk clients or engaged in condomless sex, illustrating how broader socio-economic conditions can influence behaviors with increased HIV and STI risk. Where economic recessions are coupled with shrinking public funding for health and social services ^21^, access to essential HIV testing, treatment, and prevention resources can become severely limited for sex workers.

Molecular analyses of HIV in the context of major geopolitical events can help uncover the evolutionary history and spread of the virus in relation to societal changes ^22^. These studies use viral sequencing data from individuals living with HIV to infer evolutionary relationships among viral strains and trace their spread over time and geography. A limited number of studies have linked the spread of HIV to major structural shocks and geopolitical events such as conflict ^7^, and COVID-19 restrictions ^23^, as well as slower moving, large-scale social change such as urbanization ^24^. Faria and colleagues (2014) found evidence of the role of urbanization and transport network expansion in the spread of HIV in the early 1920s from Kinshasa to other regions of sub-Saharan Africa. Others have looked at the impact of conflict, involving mass internal displacement of populations, on the spread of HIV ^25 26^. Earlier work has shown that internal displacement due to armed conflict in eastern Ukraine (Donbas region), caused a geographic redistribution of HIV lineages within the country, although it remains unknown if HIV transmission was accelerated because of conflict ^7^. Although our findings suggest that most HIV acquisition (70%) would have occurred in periods of economic decline—prior to armed conflict in eastern Ukraine—the small sample size limits our ability to draw definitive conclusions. Nonetheless, this work was designed as an exploratory analysis and aligns with other accounts describing how significant events such as economic recessions can influence HIV transmission by altering individual behaviors, the organisation of sex work, and impact HIV prevention and care programs, including in our own study population ^15 20 27 28^.

This study has several limitations. First, the cross-sectional design precludes the ability to establish temporal or causal relationships and is subject to typical biases such as recall and social desirability bias. Second, the relatively low prevalence of HIV among study participants, viral suppression due to antiretroviral therapy, and possible sample degradation likely contributed to a small sample size. While this limits the strength of our conclusions, our molecular findings were interpreted in conjunction with qualitative data, allowing for some degree of triangulation.

Although a larger sample size would be ideal, ongoing instability in Ukraine renders further data collection difficult. Alternatively, clinical records (e.g., date of first positive HIV test) could be examined to identify additional estimated times of infection and explore potential correlations with major geopolitical events—offering a feasible complement to molecular approaches. Third, ETI estimates carry an uncertainty of approximately ±1 year, which may overlap with multiple significant events in Ukraine. This complicates attribution of observed patterns to specific crises. However, most ETI estimates still fall within broader periods of economic instability, suggesting a general association between structural disruption and heightened HIV vulnerability. Finally, the generalizability of our findings is limited by the modest sample size and the focus on sex workers and their clients. Nonetheless, we argue that the broader theme—that financial pressures can influence decision-making and HIV risk—resonates beyond our study population and may apply to other marginalized groups in similar socio-economic contexts, both within and outside Ukraine.

## CONCLUSIONS

Molecular studies have the potential to support targeted HIV prevention and programming by providing important insights into the transmission dynamics of HIV. They can highlight the populations most affected by the epidemic, and improve our understanding of the relationships between social and structural factors and HIV acquisition. Molecular data adds the element of time and big events should be considered alongside HIV sequence analysis owing to their strong spatio-temporal signal. While our sample size is small, our molecular analyses add a temporal dimension to our understanding of how big events in Ukraine might have influenced patterns of HIV transmission and acquisition, leading up to the *Dynamics* Study period. Exploring connections between “upstream” big events and “downstream” HIV transmission/acquisition events (i.e. inferred time since HIV infection) can inform preparedness plans for HIV programming in anticipation of subsequent structural shocks. Using complementary data form disparate sources can also generate unique evidence for building resilient health systems that support the needs of those most vulnerable to fallout from structural shocks, including HIV. This would provide evidence for interventions rooted in human and labour rights-based approaches for populations at greatest risk of HIV during periods of geopolitical uncertainty.

## Author Contributions

François Cholette, Nicole Herpai, and Leigh M. McClarty drafted and revised the manuscript. Rupert Capiña acquired data. Olga Balakireva, Daria Pavlova, Anna Lopatenko, Paul Sandstrom, Michael Pickles, Evelyn Forget, Sharmistha Mishra, and Marissa L. Becker contributed to the conception and design of the Dynamics Study. Sharmistha Mishra and Marissa Becker critical revised and approved the final version of the manuscript.

## Institutional Review Board Statement

Ethical approval was obtained from the Health Research Ethics Board at the University of Manitoba (HS20653 [H2017:097]), Canada, the Ethical Review Committee of the Sociological Association of Ukraine, and the Committee of Medical Ethics of the L. Gromashevsky Institute of Epidemiology and Infectious Diseases at the National Academy of Medical Sciences of Ukraine.

## Informed Consent Statement

Written informed consent was obtained from study participants for the behavioural survey and biological components of the *Dynamics* Study. No identifying information was collected, but each participant was assigned an alphanumeric identification number that was used to link the behavioural and biological data. Participation lasted approximately thirty minutes and participants received an honorarium of 400 UAH (approximately $20 CAD) for their time.

## Data Availability Statement

The datasets generated and/or analysed during the current study are not publicly available due the sensitive nature of HIV molecular surveillance ^29-31^ but are available from the senior authors (Sharmistha Mishra and Marissa L. Becker) on reasonable request.

## Acknowledgments

We acknowledge the tremendous work and support of additional members of the *Dynamics* Study Team (past and present) including: Sevgi O. Aral, Tetiana Bondar, Eve Cheuk, Shajy Isac, Emma R. Lee, Christina Daniuk, Lisa Lazarus, Robert R. Lorway, Lyle McKinnon, Stephen Moses, James F. Blanchard, Maureen Murney, Huiting Ma, Nam-Mykhailo Nguien, Ani Shakarishvili, and Tatiana Tarasova. We would also like to thank the field research team and the support from DEF Group (Dnipro). This work was conducted in close partnership with non-governmental organizations (DEF Group, Ukrainian Institute for Social Research after Oleksandr Yaremenko, Alliance for Public Health in Ukraine) and governmental partners (Dnipro Oblast AIDS Centre, Center for Public Health of the Ministry of Health in Ukraine) actively involved in HIV programming. They contributed meaningfully to study design, implementation, and knowledge dissemination.

